# COVID-19 scenarios for comparing the effectiveness of age-specific vaccination regimes, exemplified for the city of Aschaffenburg (Germany)

**DOI:** 10.1101/2021.08.31.21262915

**Authors:** Tobias Krebs, Holger von Jouanne-Diedrich, Michael J. Moeckel

## Abstract

**Purpose of this report:** The purpose of this rapid communication is to illustrate the effectiveness of different vaccination regimes for controlling the number of severe and critical COVID-19 cases in the city of Aschaffenburg, Germany. Our results show that, despite numerous vaccinations in the past, further vaccinations are necessary to immunize the population and to keep the number of severe and critical cases low in the coming months. Considering that not all people can or want to receive vaccination, we compare different age-specific vaccination approaches.

**Applied Methods:** We use the agent-based epidemiological simulator Covasim for discussing the impact of different vaccination strategies. We calibrate it to reproduce the historical course of the COVID-19 pandemic in the city of Aschaffenburg, Germany; for this, we model and integrate numerous public health interventions imposed on the local population. As for some of the political actions rigorous quantification is currently not available, we fit those unknown (free) model parameters to published data on the measured epidemiological dynamics. Then we calculate the state of immunization of the population, gained through infections and vaccinations, at any time in the past, including models for time-dependent immunity decay that have been made available in Covasim. Finally, we define and compare scenarios of different vaccination regimes, especially with regard to vaccinating adolescents and providing booster vaccinations to the elderly.

**Key message:** Without further vaccinations, we expect a strong increase in severe and critical cases. In order to restrict their growth our simulations suggest that in all considered cases vaccinations of unvaccinated people is more effective than booster vaccinations for already fully vaccinated people. This applies even to vaccinations of young people who are not themselves at high risk of developing severe or critical illness. We attribute this observation to the fact that immunization of adolescents indirectly protects vulnerable age groups by preventing the spread of the virus more effectively than further immunizing other age groups. This indicates that with the pandemic ongoing, strategies focussed on minimizing individual health risks by vaccinations may no longer coincide with those needed to minimize the number of severe and critical cases.

## Background

The COVID-19 pandemic has kept the world on suspense for almost two years and rigorous public health actions have helped to control the number of hospitalizations. Initially, a new virus interacted with a mostly susceptible unprepared population. With the pandemic ongoing, a new situation has developed in August 2021: According to the German National Institute for Public Health (Robert Koch Institute, RKI), 64% of the population in Germany was at least partially vaccinated and 59% of the population was fully vaccinated. In the over-60 age group, almost one in five was still unvaccinated. Previous infections and vaccinations alike have created a significant level of specific immunization while a substantial part of the population remains susceptible to new infections. Moreover, in Germany the character of the ongoing vaccination campaigns has changed since a remarkable part of the population intentionally opts out from participating despite the availability of vaccines. This has raised new questions on how to proceed with public immunization efforts.

Since the beginning of the pandemic, epidemiologists have been using computer simulations in an attempt to meet the demands of decision-makers for scientific assessment of political options and forecasts on the development of the pandemic. A variety of models with different approaches have been adapted or newly developed for the COVID-19 pandemic (Panovska-Griffiths et al. 2021). Agent-based models have proved to be capable of representing the complexity of the pandemic in some detail, including public health interventions, such that the number of available models grew rapidly (Lorig et al. 2021). Covasim (Kerr et al. 2020) is one of these agent-based COVID-19 simulators; we adapted it for Aschaffenburg, a city in Germany with a population of about 71,000, to simulate scenarios for the city.

Covasim simulates the health status and immunization of agents in detail (Cohen et al. 2021). Agents are in a naïve state at the beginning of the simulation. Through infection, they can be either symptomatic or asymptomatic. Asymptomatic infections recover after a certain time, while symptomatic diseases start mildly and develop with certain age-specific probabilities into severe, critical or even fatal cases. For each recovered or vaccinated agent, a time dependent antibody level describes its immunity status and reduces its susceptibility to (re-) infection. Reaching its maximum a few days after infection or vaccination, a model of an exponential decay of the antibody level (Khoury et al. 2021) is assumed.

Particularly in light of a stalled vaccination campaign in Germany and the upcoming wave of infections in the autumn and winter, some scenarios are currently discussed in Germany on how to mitigate the impact of the next wave. Most under discussion are booster vaccinations for older people who have already been fully vaccinated and vaccination of adolescents aged 12 to 17.

## Methods

All our results rest on the agent-based simulation tool Covasim, which we used to simulate scenarios and compare different vaccination regimes. To exemplify our findings for a particular city, we created a synthetic population that matches statistically the real population of the German city Aschaffenburg in essential aspects, such as age structure or household composition. Importing it into Covasim, each agent represented a simplified digital twin of a real citizen. Contacts networks between the agents have been set up for four typical environments: home, school, work and free time. The simulation essentially calculates probabilities of the virus transmitting from one agent to another given existing contacts, allowing for a variety of active countermeasures. In Covasim, a stochastic procedure determines actual infection events for each day by combining specific transmission probabilities and contact networks. Thus, Covasim provides a comprehensive picture of the infection incidence in a population and depicts the temporal evolution of the epidemiological dynamics.

We generate each simulation output from an ensemble of 50 numerically equivalent implementations for the same parameter values to account for Covasim’s stochastic approach. Then, we present results as ensemble averages to reduce fluctuations and include statistical uncertainties.

Aschaffenburg is a city of some 71,000 inhabitants in the Rhine-Main metropolitan area close to Frankfurt/Main. It ranks 124 on the list of German cities by population (2020) and serves as a representative for a typical medium-sized city in Germany. It is part of the Free State of Bavaria and, hence, subject to Bavarian public health administrative orders. We integrated non-pharmaceutical (public health) and pharmaceutical (vaccinations) interventions applied in Aschaffenburg into the Covasim simulator and, whenever possible, quantitatively modelled their extent using publicly available data. We fitted the following parameters numerically to achieve consistency of retrospective simulation outputs and published health data on the real course of the pandemic.

‐ We made the base transmission probability (slightly) dependent on actions like the obligation to wear facemasks, increased hygienic barriers or enlarged average spacing of people at public places, etc.
‐ The crossover from the wild variant of COVID-19 to alpha and delta variants were modelled continuously as the simulation progresses
‐ PCR testing of the population is performed daily. We did not apply a uniform probability for selecting agents for testing; however, we ensured that infected individuals with symptomatic courses are more likely to receive testing than asymptomatic cases or uninfected individuals. Covasim sends all diagnosed agents to quarantine.
‐ At the same time, we modelled contact tracing by public health departments, which trace direct contacts of infected individuals and send them to quarantine.
‐ We included full school closures as well as hybrid teaching models with only half of the students present at school.
‐ We simulated “working from home”-arrangements by assuming a reduced number of contacts at the work level and represented private contact restrictions by a reduced number of contacts at leisure activities.
‐ During the summer months (until end of September) there is a reduced risk of infection (so-called “summer effect”), possibly because of a shift of contacts from indoor to outdoor locations. We modelled it by a correction to the risk of infection.
‐ We simulated travel during summer vacations by randomly imported infections during the vacation season.
‐ In Germany, the vaccination campaign started in December 2020. Initially, only adults received vaccine doses, subject to prioritization of both people from higher age groups or with high-risk conditions and medical staff independent of age. We approximately modelled the effect of prioritization by vaccinating elderly agents according to their age group and random adult agents as part of the medical workforce.

Procedures are described in the literature to allow for an automatic calibration process of agent-based models (Hazelbag et al. 2020). In this case, however, manual adjustment of the model parameters proved to be the best way of fitting. When calibrating the model, we fixed the free parameters of the model in such a way that the simulated curves and the provided real data of the 7-day incidence and the critical cases in the period from February 01, 2020, to July 31, 2021, visibly matched well. Then, we used the calibrated model of the pandemic as a starting point for simulating future vaccination regimes. The baseline scenario represents the obtained immunization of the population (*status quo*) as a reference.

## Data

We performed all simulations for the city of Aschaffenburg. Any data used were collected during the course of the pandemic until 3^rd^ of August 2021 and either directly included as parameters in the simulation or used as reference values for parameter fitting of the models. We downloaded data on confirmed infections and deaths in Aschaffenburg, as well as data on PCR tests and vaccinations, from the RKI. For the critical COVID-19 cases in Aschaffenburg, we consulted the DIVI intensive care unit bed registry, which documents the occupancy of the intensive care unit beds in the Aschaffenburg hospital with COVID-19 cases on a daily basis. We estimated work and leisure contacts from publicly available mobility data provided by Google. We extracted times of school closures and periods when reduced hybrid teaching models had been in place from evaluating public health orders. Other parameters such as contact tracing parameters or a reduced transmission probability in summer were estimated or used as fit parameters. Some of the relevant data, e.g. the number of PCR tests and vaccinations performed, as well as mobility data provided by Google, were not available at the level of the city of Aschaffenburg; hence, we extrapolated them from aggregated data on state (Bavaria) or national (Germany) level, assuming linear scaling with the population.

### Assumptions and simplifications

At the time of writing, the Covasim simulation software does not support by default the possibility of delivering a third vaccination to agents. Therefore, we extended the software and, in analogy, applied the already implemented model for a second vaccination to a third vaccination. With the third vaccination dose, the agents again experience a significant increase in the antibody level of their immune system to a new maximum value, similar to what has already been demonstrated in studies (Iketani et al. 2021). We note that this approach is justified to study immunity on short time scales. The model as it is may not be suitable for long-term studies, e.g. on immunity decay.

For simplicity, we assumed that the Biontech/Pfizer vaccine was used for all vaccinations.

### Scenarios

We start all scenarios from the reproduced epidemiological dynamics of the COVID-19 pandemic on 1^st^ of September 2021. Hence, we include the full immunization history of the current population as initial conditions for the subsequent evolution. For each scenario, we make different assumptions on the vaccination regime; in all cases, we simplify the calculation by administering all vaccine doses on a single, arbitrarily chosen day. We compute all simulations until 31^st^ of December 2021.

In the baseline scenario (Scenario 0), it is assumed that essentially the situation on 3^rd^ of August 2021 will persist. The future development of infections and severe and critical cases in the period up to the end of December 2021 is considered. Schools will remain permanently open during regular school hours, work attendance will remain stagnant at 86% compared to pre-pandemic levels, and leisure contacts will remain at original pre-pandemic levels, which they had reached at the time of the report. For all other parameters, such as the number of PCR tests performed, we choose the same values as in autumn 2020. Note that in the baseline scenario, the vaccination campaign has come to a full standstill and no further vaccinations take place. There is no claim that this baseline scenario complies well with reality. It serves to illustrate the current level of immunity represented in the population.

In a first scenario (Scenario 1), we study the effect of vaccination of all adolescents aged 12 to 17 years. For this, the approximately 3,500 agents in this age group receive a first dose of the vaccine from Biontech/Pfizer at the beginning of September and a second dose at the end of September, such that in total 7,000 additional vaccine doses are administered. There are no further changes to the baseline scenario. In order to ensure comparability, we us 7,000 vaccine doses in the following scenarios as well.

The second scenario (Scenario 2) addresses the case where 7,000 vaccine doses are utilised for the third vaccination of the elderly who have already received two vaccine doses (booster vaccinations). Priority is given with respect to age only, in descending order.

The third scenario (Scenario 3) combines scenarios 1 and 2 and assumes that from totally 7,000 vaccine doses 50% (i.e. 3500 doses) are applied to fully vaccinate 1750 adolescents in the age group between 12 and 17 years as described in scenario 1. The remaining 3,500 doses are used for booster vaccinations of high priority age groups (cf. scenario 2).

Finally, we consider a case complimentary to the baseline scenario: the forth scenario (Scenario 4) assumes that 7,000 vaccine doses are used to fully vaccinate previously unvaccinated people at the beginning of September 2021. The intention is to simulate the closure of remaining gaps in the protection of particularly vulnerable groups. Priority is given with respect to age only, in descending order.

## Results

For calculating the scenario simulations, we added fictitious vaccination measures described in the previous section to the calibrated model of the epidemiological evolution in Aschaffenburg. Then we continued the simulation time until 31^st^ of December 2021. Fig. 1 shows the 7-day incidence, which represents the number of confirmed COVID-19 cases per week per 100,000 inhabitants, and the daily number of treated severe and critical cases for the scenarios considered from the beginning of September to the end of December 2021.

**Fig. 1:**
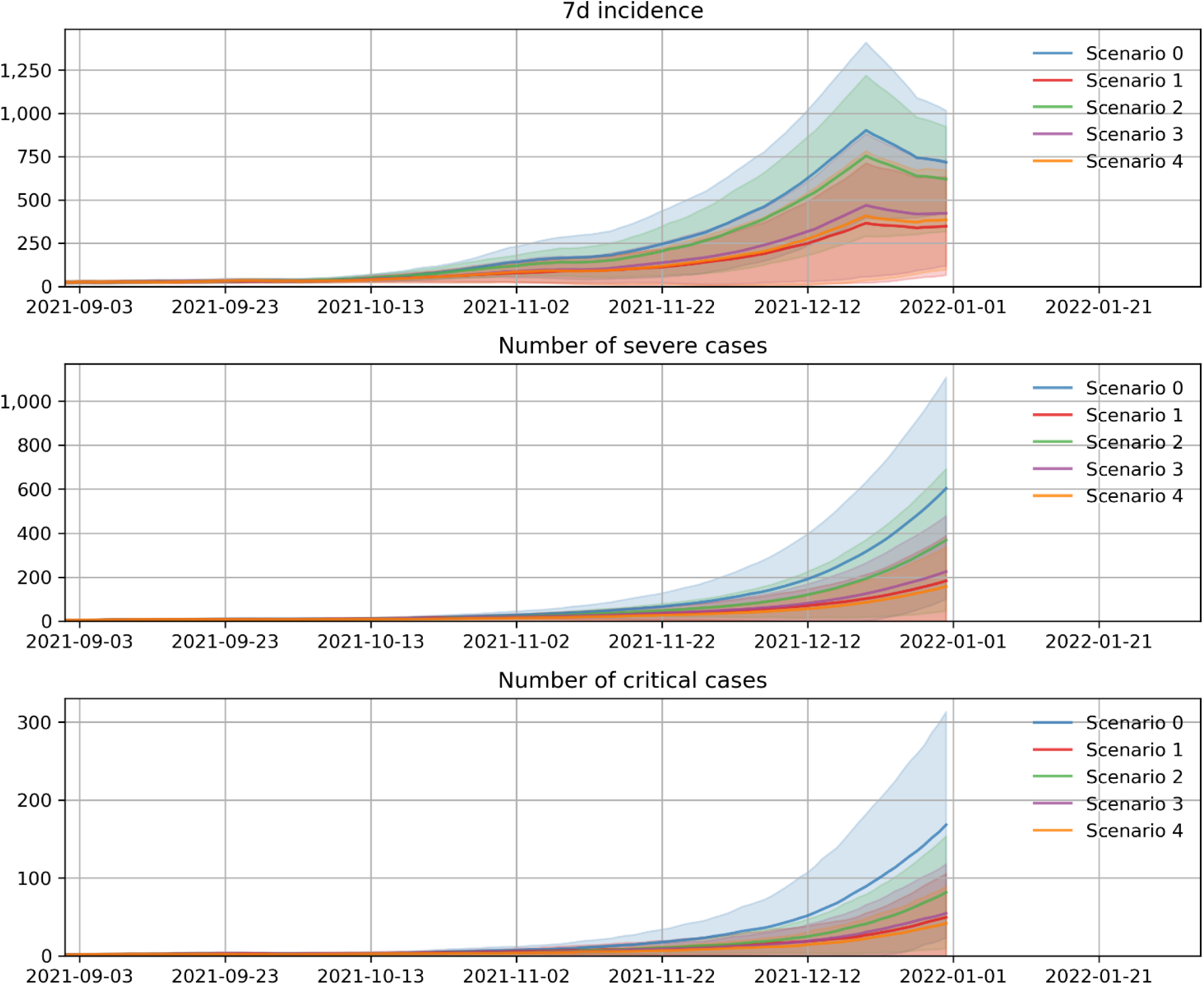
7-day incidence per 100,000 inhabitants and the absolute number of severe and critical cases for the population of 71,000 inhabitants of Aschaffenburg: the baseline scenario (scenario 0) refers to no further vaccinations; the vaccination of unvaccinated young people (scenario 1) suggests substantial contributions to pandemic control.

The shape of the curves is very similar in all scenarios considered. Both the 7-day incidence and the number of severe and critical cases show an exponential increase until Christmas. We relate the drop in infections around Christmas, among other aspects, to school vacations with a reduced number of contacts among pupils. Due to incubation and hospitalization times this kink is not seen in the number of severe or critical cases. Note that our results show ensemble averages of 100 numerical realizations for identical parameter settings; shaded areas in related colours indicate the related statistical uncertainties.

The baseline scenario (Scenario 0, no further vaccinations) shows, as expected, the highest 7 days incidence and the largest number of severe and critical cases. Hence, any vaccination strategy modelled in these scenarios reduces the impact of the pandemic substantially below the baseline scenario.

However, the number of prevented infections, severe and critical cases depends on the chosen vaccination strategy. Optimal results on limiting severe and critical cases is seen in scenario 4 (closing of vaccination gaps), i.e. by continuing with an age-dependent priority scheme. This scheme, however, may not be feasible in real life.

Therefore, we stress that we observe a very similar reduction of the number of severe and critical cases in scenario 1, which represents a completely different vaccination strategy of immunizing younger unvaccinated people. This scenario ranks second best in reducing the number of severe and critical cases and even outperforms all other strategies on the 7-day incidence.

All scenarios in which unvaccinated people are vaccinated (Scenario 1, 3, 4) have a significantly more positive effect on pandemic development than restricting to booster vaccinations of older people who are already fully vaccinated (Scenario 2). In accordance, the mixed scenario consisting of vaccination of adolescents and booster vaccinations for the elderly shows a comparatively high effectiveness in reducing severe and critical cases.

In conclusion, it is particularly remarkable that the vaccination of adolescents in scenario 1 shows a similar positive effect as the vaccination of the previously unvaccinated elderly in scenario 4.

## Discussion

Assuming that there will be no further lockdown to reduce contacts and that the number of new vaccinations will remain at a low level, the number of infections and, in particular, the number of hospitalizations is expected to rise significantly again in the autumn and winter of 2021. Severe and critical cases would primarily affect both unvaccinated elderly persons and vaccinated elderly persons whose immunization has waned over the months.

Adolescents aged 12 to 17 are at very low risk for a severe or critical course of infection. However, high vaccination coverage in this age group can make a significant contribution to reducing severe and critical cases. In particular, with open schools as well as leisure contacts on pre-pandemic levels, our simulations predict an increasing importance of this age group for the spread of infections, if vaccination rates among students remain low. We conclude that the immunization of adolescents can contribute to prevent potential infection chains and thus reduce the virus exposure of unprotected high-risk individuals of other age groups. While this indirect effect of protecting other members of society exists for all vaccinated people, it is most effective in groups with large contact networks, e.g. for schoolchildren. Seen from this perspective, vaccinating an older person with a small contact network primarily reduces his or her personal risk, which reduces the number of severe or critical cases in accordance to the personal risk for severe illness of the vaccinated person. The personal advantage of vaccination for an adolescent is, due to his or her reduced personal risk, smaller. Hence, reductions in the number of severe or critical cases are not primarily originating from the prevention of infections or serious cases among the vaccinated adolescents. Instead, vaccinating an adolescent reduces the risk for every person in the population by a small amount. Especially unvaccinated members of high-risk groups benefit most from this suppression of the epidemiological dynamics. Because of the current immunity of the population reached by previous vaccinations and the higher number of contacts of adolescents the aggregated collective effects of vaccinating younger age groups, appear similar to those of directly vaccinating high-risk groups, as our simulations suggest.

Older, vaccinated individuals experience slowly waning immunization. They can considerably reduce their personal risk of infection by receiving a booster vaccination. Thus, severe and critical courses of infections can also be prevented by administering a third dose. However, based on our simulation results, we point out that third-dose vaccination alone will not be sufficient to keep hospitalizations low this autumn and winter. We attribute this partly to the comparatively large number of unvaccinated individuals in the most vulnerable age groups of the population who either voluntarily opt out of vaccination campaigns or do not receive vaccination for medical reasons.

## Limitations

### Special limitation of our study

Immunity against COVID-19 consists of both an antibody and a T-cell response. While antibody levels decay after some time, T-cell related immunity, which is not explicitly part of immunity decay models, may persist longer. We note that if possible long-term immunity effects have been neglected in our simulations, the total immunity of vaccinated people will be underestimated. This however, implies that qualitative results will not be affected and general conclusions remain valid. Nonetheless, we restrict ourselves not to extrapolate the immunity status of vaccinated people too far into the future and limit all simulations to the year 2021.

In comparing vaccination regimens, we focussed on benefits for the society, i.e. aiming at minimizing the total number of severe and critical courses of infection. We could not and did not include aspects of individual risk assessment. In particular, it has not been discussed to which extend the low likelihood of contracting a severe or critical course of COVID-19 among young people justifies the acceptance of known or possibly unknown risks involved in vaccination on an individual basis. Therefore, the authors do not intend to advise or discourage people to seek vaccination against COVID-19 individually. Nonetheless, we stress that our observations suggest that for controlling the COVID-19 pandemic society profits from further immunization efforts in a way that is not always clearly linked to individual health risks.

The results of this report are to be considered preliminary and may change with further data and findings collected in the future. In particular, only findings and local data for the City of Aschaffenburg have been included. Little data is available on the actual implementation of public health orders. Therefore, we had to refer to plausible assumptions or fitted parameters for modelling some public health interventions. The stability of our results with respect to changes in those assumptions has not been established in all cases, due to time constraints. In principle, modified assumptions may have some influence on the simulations performed.

### General limitations of epidemiological simulations

The greatest uncertainty of our simulation lies in the actual human behaviour. The degree of compliance is difficult to measure, but has a non-negligible impact on the evolution of the pandemic. Past data was used to calibrate, among other things, a baseline probability of transmission of the virus in a contact between two agents. This probability includes aspects such as wearing masks, following distance rules, and the like. If the average behaviour changes in this regard, it is difficult to account for. In particular, we did not try to predict any behavioural changes from September until December 2021 but kept all related simulation parameters constant.

## Conclusions

We have simulated the 7-day incidence and the absolute number of severe and critical COVID-19 cases using Covasim for comparing different fictitious vaccination strategies applied to the population of the German city of Aschaffenburg. As expected, direct vaccination of people from high-risk groups who have not previously been vaccinated is particularly effective. The scenarios considered also show that vaccination of younger population groups can significantly increase the overall protection of the population. From a societal perspective, a high vaccination rate in all age groups is desirable. Booster vaccinations for fully vaccinated people increase their individual protection and, in consequence, reduce the total number of severe and critical cases. However, they do not achieve the greatest collective benefit in terms of controlling the number of severe and critical cases in the future. We conclude that at the current stage of the COVID-19 pandemic further vaccination efforts are helpful and necessary to limit the number of severe and critical cases. Yet vaccination strategies solely based on individual risk assessment and an individualized perspective of personal risk may not be the only and not necessarily the best option to control the pandemic.

## Data Availability

Only publicly available data has been used

## Acknowledgements

The authors gratefully acknowledge funding by the EpiL**AB**^KI^ project through the Bavarian Programme for Applied Research and Development at Universities of Applied Sciences (6^th^ funding period).

## References

Cohen Jamie A.; Stuart Robyn M.; Núñez Rafael C.; Wagner, Bradley; Chang, Stewart; Rosenfeld, Katherine et al. (2021): Mechanistic modeling of SARS-CoV-2 immune memory, variants, and vaccines. In: medRxiv, 2021.05.31.21258018. DOI: 10.1101/2021.05.31.21258018.

Hazelbag C. Marijn Dushoff, Jonathan; Dominic, Emanuel M.; Mthombothi Zinhle E.; Delva, Wim (2020): Calibration of individual-based models to epidemiological data: A systematic review. In: PLOS Computational Biology 16 (5), e1007893. DOI: 10.1371/journal.pcbi.1007893.

Iketani Sho; Liu, Lihong; Nair, Manoj S.; Mohri, Hiroshi; Wang, Maple; Huang, Yaoxing; Ho David D. (2021): A third COVID-19 vaccine shot markedly boosts neutralizing antibody potency and breadth. In: medRxiv, 2021.08.11.21261670. DOI: 10.1101/2021.08.11.21261670.

Kerr Cliff C.; Stuart Robyn M.; Mistry, Dina; Abeysuriya Romesh G.; Rosenfeld, Katherine; Hart Gregory R. et al. (2020): Covasim: an agent-based model of COVID-19 dynamics and interventions. In: PLOS Computational Biology. DOI: 10.1101/2020.05.10.20097469.

Khoury, David S.; Cromer, Deborah; Reynaldi, Arnold; Schlub Timothy E.; Wheatley Adam K.; Juno Jennifer A. et al. (2021): Neutralizing antibody levels are highly predictive of immune protection from symptomatic SARS-CoV-2 infection. In: Nat Med 27 (7), S. 1205–1211. DOI: 10.1038/s41591-021-01377-8.

Lorig, Fabian Johansson, Emil; Davidsson Paul (2021): Agent-Based Social Simulation of the COVID-19 Pandemic: A Systematic Review. In: JASSS 24 (3). DOI: 10.18564/jasss.4601.

Panovska-Griffiths, J.; Kerr, C. C.; Waites, W.; Stuart, R. M. (2021): Mathematical modeling as a tool for policy decision making: Applications to the COVID-19 pandemic. In: Data Science: Theory and Applications, Bd. 44: Elsevier (Handbook of Statistics), S. 291–326.

